# The impact of lidocaine plaster prescribing reduction strategies: a comparison of two national health services in Europe

**DOI:** 10.1101/2023.02.23.23286366

**Authors:** Molly Mattsson, Fiona Boland, Ciara Kirke, Michelle Flood, Emma Wallace, Mary E Walsh, Derek Corrigan, Tom Fahey, Richard Croker, Sebastian C J Bacon, Peter Inglesby, David Evans, Ben Goldacre, Brian MacKenna, Frank Moriarty

**Affiliations:** School of Pharmacy and Biomolecular Sciences, Royal College of Surgeons in Ireland (RCSI) University of Medicine and Health Sciences, Dublin, Ireland; Data Science Centre, RCSI University of Medicine and Health Sciences, Dublin, Ireland; National Medication Safety Programme, HSE National Quality and Patient Safety Directorate, Dublin, Ireland; Department of General Practice, University College Cork, Cork, Ireland; School of Public Health, Physiotherapy and Sports Science, University College Dublin, Ireland; FutureNeuro Research Centre (eHealth Group), RCSI University of Medicine and Health Sciences, Dublin, Ireland; Department of General Practice, RCSI University of Medicine and Health Sciences, Dublin, Ireland; Bennett Institute for Applied Data Science, Nuffield Department of Primary Care Health Sciences, University of Oxford, Oxford, United Kingdom

## Abstract

**Introduction:** In 2017, two distinct interventions were implemented in Ireland and England to reduce prescribing of lidocaine medicated plasters. In Ireland, restrictions on reimbursement were introduced. In England, updated guidance on items not to routinely prescribe in primary care, including lidocaine plasters, was published. This study aims to compare how the interventions impacted prescribing of lidocaine plasters in these countries.

**Methods:** We conducted an interrupted time series study using general practice data. For Ireland, monthly dispensing data (2015-2019) from the means-tested General Medical Services scheme was used. For England, data covered all patients. Outcomes were the rate of dispensings, quantity and costs of lidocaine plasters, and we modelled level and trend changes from the first full month of the policy/guidance change.

**Findings:** Ireland had higher rates of lidocaine dispensings compared to England throughout the study period; this was 15.22/1,000 population immediately pre-intervention, and there was equivalent to a 97.2% immediate reduction following the intervention. In England, the immediate pre-intervention dispensing rate was 0.36/1,000, with an immediate reduction of 0.0251/1,000 (a 5.8% decrease), followed by a small but significant decrease in the monthly trend relative to the pre-intervention trend of 0.0057 per month.

**Interpretation:** Among two different interventions aiming to decrease low-value lidocaine plaster prescribing, there was a substantially larger impact in Ireland of reimbursement restriction compared to issuing guidance in England. However, this is in the context of much higher baseline rates of use in Ireland compared to England.

**Funding:** This study was funded by the Health Research Board (SDAP-2019-023).

## Background

Regardless of healthcare system, the resources available for healthcare are limited compared with demand, and all healthcare systems consequently employ mechanisms to prioritise finite healthcare resources to maximise health benefits.^1^ One integral part of healthcare delivery is the provision of medicines, which accounts for a significant proportion of overall health expenditure in most countries. In 2019, spending on retail pharmaceuticals (excluding those used during hospital treatment) accounted for one-sixth of overall health care expenditure in Organisation for Economic Co-operation and Development (OECD) countries and represented the third largest component of health spending after inpatient and outpatient care^2^.

Rational use of medicines requires that “patients receive medications appropriate to their clinical needs, in doses that meet their own individual requirements, for an adequate period of time, and at the lowest cost to them and their community”^3^. In contrast, irrational use of medicines is a major problem worldwide, with the World Health Organization (WHO) estimating that more than half of all medicines are prescribed, dispensed or sold inappropriately, and that half of all patients fail to take them correctly^4^. Low-value care, the use of health services whose harms or costs exceed their benefits, is also a significant issue that contributes to wasted healthcare resources^5^. Various strategies exist to promote rational prescribing, aimed at both patients and prescribers, and ensure safe, effective, and cost-effective medicines use. These strategies can be grouped broadly as targeted or system-oriented approaches, with targeted approaches comprising educational and managerial interventions and system-oriented strategies including regulatory and economic interventions^6^.

Both the National Health Service (NHS) in England and the Health Service Executive (HSE) in Ireland identified prescribing of lidocaine 5% medicated plasters (Versatis®) as a target for prescribing reduction measures. This medicinal product’s licensed indication is for the treatment of post-herpetic neuralgia (PHN) only. However, it had been prescribed and dispensed in volumes exceeding the likely prevalence of PHN, indicating off-label use. In Ireland, the Medicines Management Programme (MMP) was established in 2013, with the aim to provide sustained national leadership relating to issues such as the quality of the medicines management process, access to medicines and overall expenditure on medicines^7^. In March 2017, the MMP published a Prescribing and Cost Guidance document on the lidocaine 5% medicated plasters^8^. Following publication of these guidelines, the HSE introduced changes to the reimbursement of lidocaine plasters, introduced in two stages. From 1^st^ September 2017 prescribers are required to apply through an online reimbursement applications system for all new patients, indicating the antiviral that was prescribed for the herpes zoster infection, and the date it was prescribed. From 1^st^ December 2017 this was extended to pre-existing patients in receipt of the medication prior to September 2017.

Similarly in March 2017 NHS England announced a programme to tackle “low value medicines^9^, which subsequently became guidance on *items which should not routinely be prescribed in primary care*. This included lidocaine plasters, which was classified as “an item of low clinical effectiveness, where there is a lack of robust evidence of clinical effectiveness or there are significant safety concerns”^10^. This guidance advised that prescribers in primary care should not initiate lidocaine plasters for any new patients (unless patients have been treated in line with NICE CG173 “Neuropathic pain in adults: pharmacological management in non-specialist settings”, but are still experiencing PHN), that prescribers should be supported in deprescribing lidocaine plasters in all patients, and that if there is a clinical need for lidocaine plasters to be prescribed in primary care, this should be undertaken in a cooperation arrangement with a multi-disciplinary team^10^.

This study aims to describe and compare how the policy and guidance changes have impacted prescribing of lidocaine plasters in the two countries.

## Methods

We conducted an interrupted time series study using segmented regression analysis to assess the change in prescribing rate following the introduction of guidance and policy changes. Interrupted time series studies of policy interventions can be analysed using segmented regression, allowing for the change in level and trend of an outcome following an intervention to be evaluated^11^. The Strengthening the Reporting of Observational studies in Epidemiology (STROBE) guidelines are used in reporting this study^12^. This study was approved by the RCSI University of Medicine and Health Sciences Human Research Ethics Committee (REC202201015).

### HSE data

The Primary Care Reimbursement Service (PCRS) is the section within the HSE which administers community drug schemes in Ireland, including the GMS scheme. Eligibility for this scheme is based on age and income and covers approximately 32% of the population, and therefore eligible persons tend to be more socioeconomically deprived than the general population^13^. However, the scheme does cover the vast majority of adults aged 70 years and over, and the data provides complete information on prescribed medications that are dispensed to eligible people. For this scheme, pharmacies transmit claims for medications to the PCRS at the end of each month for reimbursement. The pharmacy claims database from the PCRS contains records of prescribed medications which were dispensed to individuals eligible for community drug schemes. As data is not publicly available, a request was submitted to the PCRS in line with their information requests policy, to obtain data aggregated at local health office (LHO) level. The data contains drug information (WHO Anatomical Therapeutic Chemical (ATC) code, strength, defined daily dosage (DDD), and product information), quantity dispensed, month of dispensing, and cost.

### NHS data

The NHS openly publishes GP prescribing data every month and it is available from the OpenPrescribing.net platform, which provides monthly statistics of prescribing of different medications aggregated at the level of GP practices for all practices in England^14^. The data relates to NHS prescriptions issued by general practices in England (by any practice prescribing staff) and dispensed in any community pharmacy in the UK. Prescribed products are coded based on their British National Formulary (BNF) classification. The monthly prescribing datasets contain one row for each different medication and dose in each prescribing organisation in NHS primary care in England, describing the number of items and the total cost. These data are sourced from community pharmacy claims data and, therefore, contain all items that were dispensed. The items variable within the data corresponds to the number of items of each prescribed product that was dispensed in the specified month. This provides comparable rates to the ‘number of prescription dispensings’ indicator as defined in the PCRS data. The data is available at practice-level and was aggregated to clinical commissioning group (CCG) level, an NHS administrative region, to allow for equivalent analysis to the PCRS data.

### Analysis

Firstly, descriptive statistics were used to summarise dispensings, quantity, and costs of lidocaine plasters on a monthly basis in the two countries. We also summarised dispensings of alternatives recommended by the HSE Medicines Management Programme using HSE data^8^. These were plotted to allow visual inspection of trends in outcomes in relation to the policy and guideline changes. A segmented regression model was fitted separately on the HSE and NHS data to assess the change in prescribing rates following the intervention affecting lidocaine plasters in each country (i.e., introduction of reimbursement changes/guidelines).

We parameterised each segmented regression model to estimate four elements:

i. the rate at the beginning of the study period (i.e., the model intercept),
ii. the trend prior to the intervention,
iii. immediate change in rate from pre to post intervention, and
iv. change in trend over time from pre to post intervention.

The analysis included monthly data from 2015 to 2019, allowing for well in excess of the recommended twelve time points before and after an interruption. As Ireland introduced the intervention in stages, with the reimbursement application initially required for only patients newly initiating lidocaine plasters (1^st^ September 2017, intervention 1) before being extended to all patients (1^st^ December 2017, intervention 2),, the effects of both interventions were included. For England, August 2017 was considered the first month post-intervention, as the guidance was first published in July 2017 as part of a consultation. Analyses were conducted using Stata version 17, and statistical significance was assumed at p<0.05. For the time-series analysis, the XTITSA command was used, which allows for analysis of panel data at LHO/CCG level^15^.

## Results

### Lidocaine patches

In Ireland, there were 193,486 individual dispensings for lidocaine plasters in 2015, reducing to 21,886 in 2019. The GMS expenditure similarly decreased during the study period, peaking at €27.4 million in 2016 compared to €2.7 million in 2019. In England, a slight reduction in dispensings was noted during the study period, with dispensings peaking at 258,574 in 2017, and decreasing to 219,177 in 2019. Costs decreased from £19,428,950 in 2017, to £16,211,567 in 2019. Table 1 outlines the year-by-year dispensings, quantity, and costs for lidocaine patches for Ireland and England.

**Table 1.** Year-by-year dispensings, quantity, and costs for Lidocaine patches by country.

| Lidocaine 5% medicated plasters |  |  |  |  |  |  |
| --- | --- | --- | --- | --- | --- | --- |
|  | Ireland |  |  | England |  |  |
|  | Dispensings | Quantity | Cost (€) | Dispensings | Quantity | Cost (£) |
| **2015** | 193,486 | 6,033,805 | 24,262,047 | 223,541 | 6,873,093 | 17,289,944 |
| **2016** | 249,075 | 7,795,933 | 27,321,197 | 254,205 | 7,834,000 | 18,909,296 |
| **2017** | 235,437 | 7,497,794 | 25,138,152 | 258,574 | 8,085,800 | 19,428,950 |
| **2018** | 16,332 | 602,345 | 2,004,115 | 239,556 | 7,586,363 | 17,850,706 |
| **2019** | 21,886 | 826,653 | 2,746,972 | 219,177 | 7,007,670 | 16,211,567 |

Ireland had higher rates of dispensings compared to England throughout the study period. In Ireland, the rate per 1,000 eligible GMS population was 8.15 dispensings in January 2015, 15.22 in August 2017, and 1.15 in December 2019. In England, the mean rate per 1,000 NHS population was 0.28 dispensings in January 2015, 0.36 in July 2017, and 0.30 in December 2019. See Table 2 and Figure 1 for dispensings rate per 1,000 eligible population for Ireland and England.

**Table 2.** Year-by-year rates of Lidocaine patch dispensings by country.

| Lidocaine 5% medicated plasters |  |  |  |  |
| --- | --- | --- | --- | --- |
|  | Ireland |  | England |  |
|  | Rate of dispensings | 95% CI | Rate of dispensings | 95% CI |
| **2015** | 9.58 | 8.97-10.18 | 0.31 | 0.30-0.33 |
| **2016** | 12.33 | 11.85-12.80 | 0.35 | 0.35-0.36 |
| **2017** | 11.65 | 9.05-14.26 | 0.36 | 0.35-0.37 |
| **2018** | 0.81 | 0.71-0.90 | 0.33 | 0.33-0.34 |
| **2019** | 1.08 | 1.05-1.12 | 0.31 | 0.30-0.31 |

**Figure 1.**
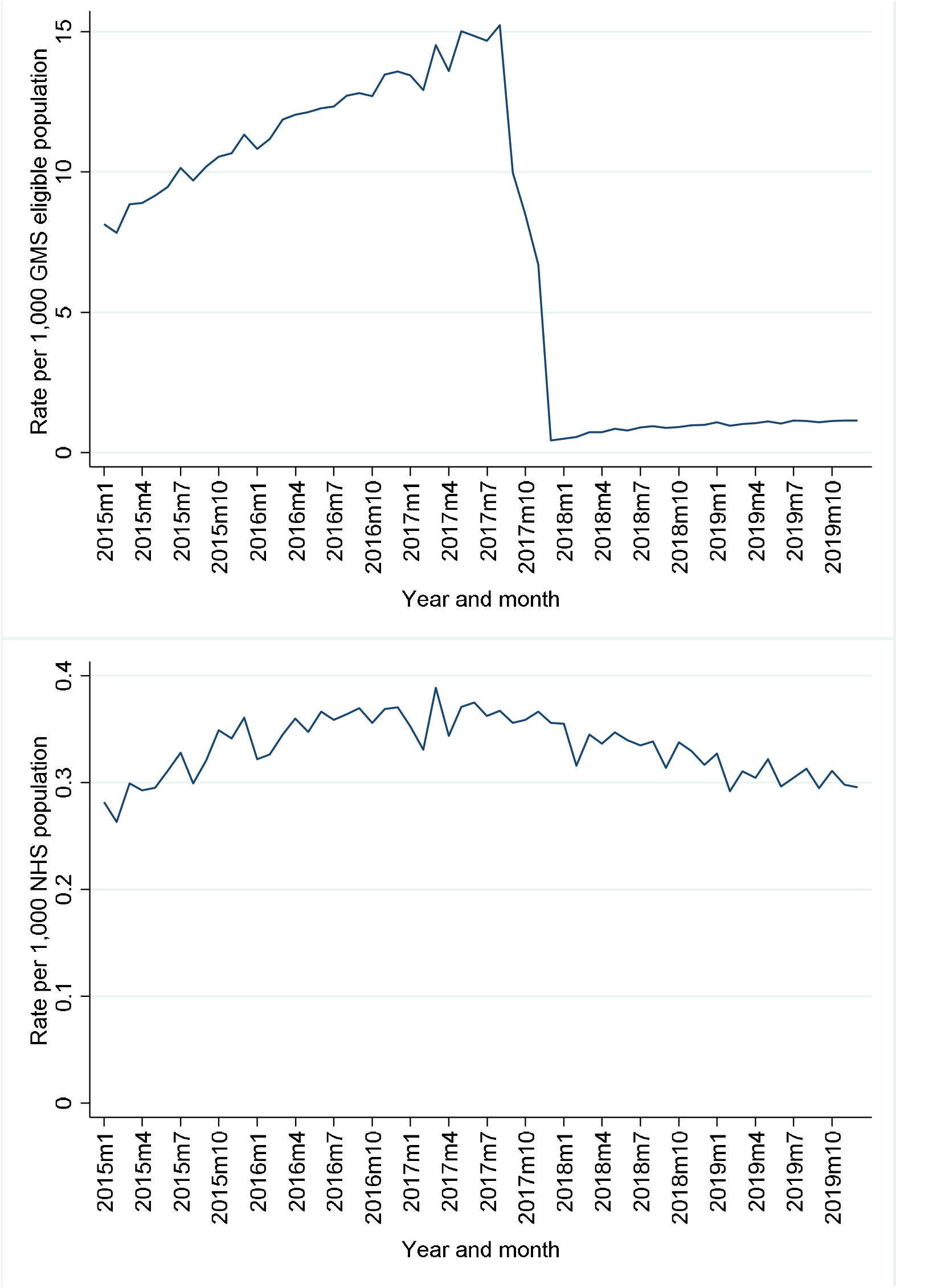
Lidocaine patches dispensings per 1,000 eligible population for Ireland (top) and England (bottom)

### Impact of policy and guidance changes

Interrupted time-series regression results for Ireland and England are outlined in Table 3. For Ireland, the dispensing rate per 1,000 GMS eligible population was estimated at 8.59 in January 2015, and appeared to increase monthly prior to September 2017, by 0.23 on average. In the first month post intervention 1, there appeared to be a statistically significant decrease in the level of 5.54 (i.e., change in rate from August 2017 to September 2017), followed by a statistically significant average decrease in the monthly trend, relative to the pre-intervention trend, of 1.93 per month. At the second intervention-point (December 2017) there was an immediate significant decrease in rate of dispensings of 4.48 (i.e., November 2017 to December 2017). After intervention 1 there was a decreasing monthly trend, of 1.7, however from December 2017, after intervention 2, there was evidence of a 0.02 monthly increase in trend. Overall, there was a decrease of 15.14 (95%CI 14.76 to 15.53) in the rate per 1,000 GMS eligible population between August (month prior to first intervention) and December 2017 (month following the second intervention), with the trend from December 2017 (after interventions) 0.2 (95%CI 0.18-0.23) lower than that prior to any interventions. This equates to a 97.3 percent (95%CI 94.8 to 99.8) reduction in the dispensing rate post-relative to pre-intervention.

**Table 3.** Dispensings time-series results for Ireland (GMS scheme) and England (NHS)

| **Rate of lidocaine dispensings per 1,000 population** | **Coefficient** | **95% CI** | **P-value** |
| --- | --- | --- | --- |
| **Ireland** |  |  |  |
| Pre-Sep 2017 monthly trend | 0.23 | 0.21 to 0.24 | <0.001 |
| Sep 2017 immediate post intervention change | -5.54 | -6.26 to -4.83 | <0.001 |
| Sep 2017 – Nov 2017 trend change (versus pre-Sep 2017) | -1.93 | -2.44 to -1.42 | <0.001 |
| Dec 2017 immediate post intervention change | -4.48 | -5.63 to -3.34 | <0.001 |
| Dec 2017 – Dec 2019 trend change (versus pre-Sep 2017) | 1.73 | 1.22 to 2.24 | <0.001 |
| Jan 2015 dispensing rate (baseline) | 8.59 | 7.7 to 9.48 | <0.001 |
| **Postintervention linear trends** |  |  |  |
| Sep 2017 – Nov 2017 | -1.70 | -2.22 to -1.19 | <0.001 |
| Dec 2017 – Dec 2019 | 0.02 | 0.005 to 0.04 | 0.016 |
| **England** |  |  |  |
| Pre-Aug 2017 monthly trend | 0.0032 | 0.0027 to 0.0036 | <0.001 |
| Aug 2017 immediate post intervention change | -0.0251 | -0.036 to -0.0142 | <0.001 |
| Aug 2017 – Dec 2019 trend change | -0.0057 | -0.0064 to -0.0051 | <0.001 |
| Jan 2015 dispensing rate (baseline) | 0.3566 | 0.2764 to 0.4367 | <0.001 |
| **Postintervention linear trends** |  |  |  |
| Aug 2017 – Dec 2019 | -0.0026 | -0.0031 to -0.0021 | <0.001 |

In England, the dispensings rate per 1,000 NHS population was estimated at 0.36 in January 2015 and appeared to increase every month prior to August 2017 by 0.0032. In the first month post the intervention (August 2017), there was a statistically significant decrease in the level of 0.0251, equating to a 5.8 percent (95%CI 3.3 to 8.4) reduction in the dispensing rate post-relative to pre-intervention. This was followed by a statistically significant decrease in the monthly trend relative to the pre-intervention trend of 0.0057 per month, The estimated trend after the intervention decreased monthly at a rate of 0.0026. Figures 2a and 2b provide a visual representation of the time-series analysis.

**Figure 2.**
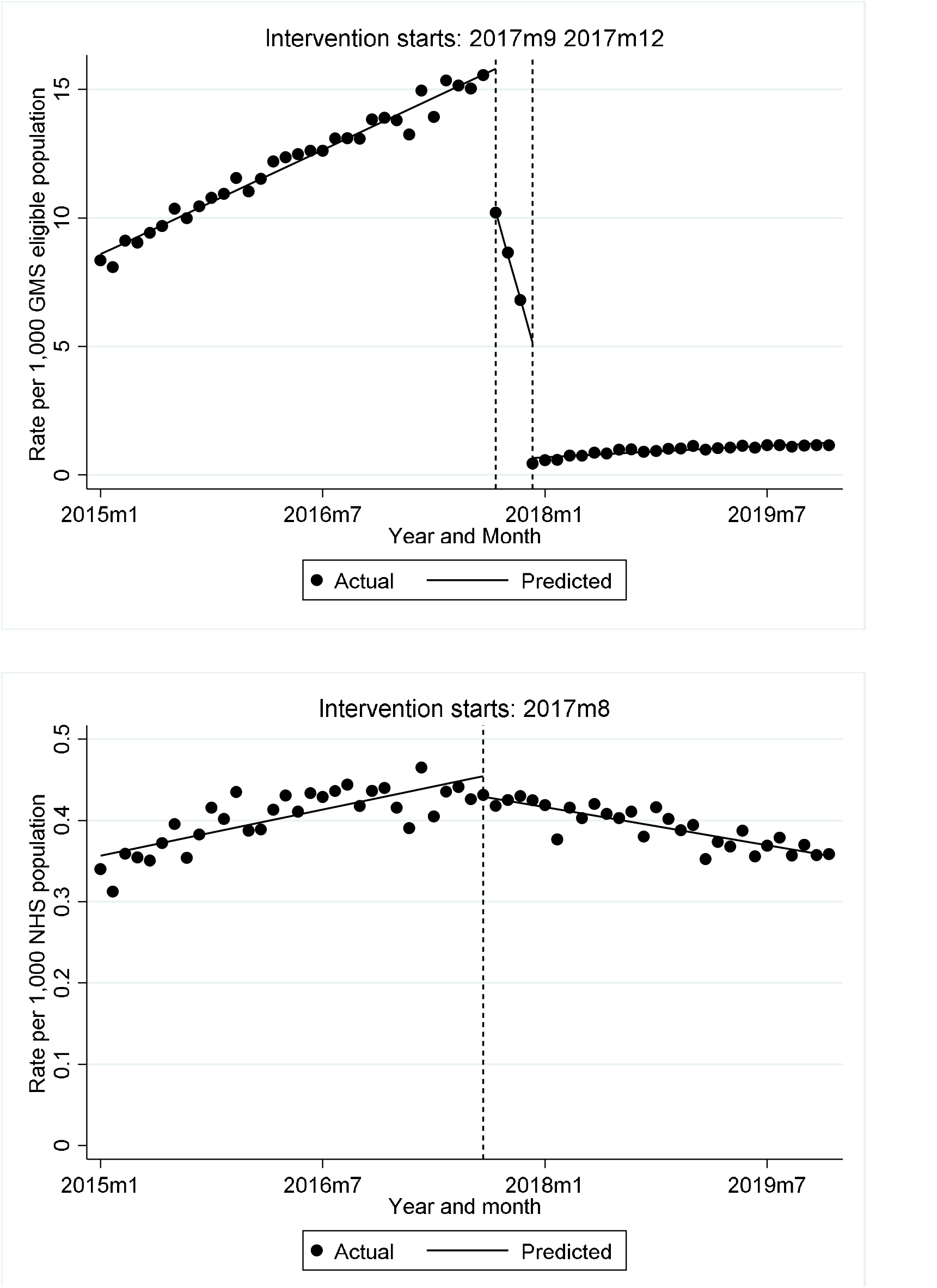
Time-series of dispensing rate of lidocaine patches per 1,000 for Ireland (top) and England (bottom)

For the quantity of lidocaine plasters, similar results as for the dispensings were found for both Ireland and England (supplementary tables 1a and 1b). In Ireland, an overall decrease of 472.45 (95%CI 459.31 to 485.58) in the rate per 1,000 GMS eligible population was seen between September and December 2017, with the trend from December 6.15 (95%CI 5.31 to 6.98) lower than that pre-September. In England, August 2017 saw a level change of -0.62 (95%CI -0.94 to -0.3), followed by a trend change of -0.17 (95%CI -0.19 to -0.15) compared to the pre-intervention trend. Similar changes in the cost of lidocaine plaster dispensing (supplementary tables 2a and 2b).

### Topical alternatives in Ireland

For the topical alternatives recommended as part of the intervention in Ireland, the overall level change in the rate of capsaicin dispensings per 1,000 GMS eligible population between September and December 2017 was 0.40 (95% CI 0.37 to 0.44), with the trend from December -0.02 (95%CI -0.022 to -0.018) lower than that pre-September. For topical NSAIDs, the overall level change in the rate per 1,000 GMS eligible population between September and December 2017 was 0.61 (95% CI 0.24 to 0.98), with the trend from December 0.03 (95%CI 0.01 to 0.06) higher than that pre-September. No significant change in level or trend was observed for topical diclofenac specifically following the intervention. Supplementary figures 3a-c provide a visual representation of the time-series analysis.

## Discussion

The findings of this study highlight the vastly different effects of the introduction of reimbursement restrictions in Ireland and guidelines in England. However, this is in the context of a much higher baseline rate of use in Ireland compared to England. In Ireland, the dispensing rate prior to the intervention was over 15 per 1,000 GMS population. The change to reimbursement had a dramatic effect on lidocaine use, with a 97.3 percent reduction in the dispensing rate of lidocaine medicated plasters post-intervention compared to before. In England, only a small decrease of 5.8 percent was seen after the guidance changes, however the dispensing rate prior to the intervention never went above 0.5 per 1,000 population. In terms of the recommended alternatives, advice accompanying the reimbursement change in Ireland may have influenced topical capsaicin use in the immediate term, however this quickly reduced.

The substantial difference in effect may be attributable to the difference in baseline rates, which may indicate that in England, lidocaine prescribing was already at a largely appropriate level prior to the introduction of the guidelines, whereas in Ireland lidocaine may have been overprescribed. Secondly, this could be attributable to the different types of intervention, i.e., a restriction on reimbursement requiring an individual application for a patient to continue to be covered for it (or alternatively paying out-of-pocket which for many patients is likely unaffordable), versus guidance on low-value care without any stringent restrictions, penalties, or incentives. A previous systematic review on the effect of formulary restrictions on drug and health care resource utilisation and economic outcomes, as well as patient outcomes, found that more than 90% of the included studies showed lower drug utilisation after introduction of reimbursement changes. However, when considering all outcomes, around half were negative in direction or unfavourable, compared to around 40% which were positive in direction or favourable, which shows that these types of interventions may have unintended consequences despite lower drug utilisation and medication cost savings^16^. As only aggregate data was analysed for this study, it was not possible to assess the impact of the interventions on individual patients, including switching behaviours and utilisation of other types of analgesia, and further research is warranted. The authors will investigate these issues further in a separate study^17^.

Low-value care and treatments have been receiving increased attention in recent years, and there has been a rapid growth in studies of interventions that target low-value care. A systematic review of measures used to assess the impact of interventions to reduce low-value care found that most published studies (68%) focused on reductions in utilisation rather than on potentially more clinically meaningful measures, such as improvements in appropriateness or patient outcomes^18^. In England, research on the trends and variation in prescribing of these low-priority treatments prior to the introduction of the guidelines found that prescribing was extensive but varied widely by treatment, geographic area and individual practice, with the proportion of patients aged 65 year and over at practice level, as well as CCG, strongly associated with low-value prescribing^19^.

An evaluation of the NHS guidance on items which should not routinely be prescribed in primary care has shown that although there was a reduction in overall use of the targeted medications, that reduction was in line with the existing downward trend, with no change either after the announcement of the consultation on the scheme (July 2017) or publication of the subsequent consultation report (November 2017)^20^. Previous research on the implementation of new antibiotic prescribing guidelines for urinary tract infection in NHS England primary care suggest that the variation between CCG may be substantial, and that there is strong evidence suggesting that CCGs with minimal prescribing change post the introduction of new guidance did less to implement changes compared to CCGs that saw positive change^21^. In Ireland, recent research has shown that subsequent to the change in reimbursement, the prescribing of lidocaine plasters significantly decreased, with the annual General Medical Services (GMS) expenditure on lidocaine plasters decreasing from €27 million in 2016 to just over €2 million in 2018^22^.

Considering implementation of guidance as an intervention, a systematic review of the evidence to practice gap for complex interventions in primary care found that success is influenced by factors related to the external context (e.g. policies and infrastructure), organisation (e.g. culture and resources), individual (e.g. competency), and intervention (e.g. evidence of benefit and ease of use)^23^. A recent scoping review of strategies for de-implementation of low-value care found that with a few exceptions similar strategies are used for de-implementation and implementation^24^.

One explanation for the substantial difference in prescribing rates between the two countries may be explained by the difference in study populations and access to healthcare. While the NHS data includes all prescriptions dispensed nationally, the HSE data is restricted to individuals eligible for the means-tested GMS scheme, where there is an over-representation of older adults, and people with lower socioeconomic status, both of which groups experience higher prevalence of chronic pain. In older adults, pain is one of the most widely cited symptoms underlying disability among older adults, and with a wide range of estimated prevalence between 25 and 75%^25, 26^. Similarly, population studies reliably show that the prevalence of chronic pain is inversely related to socio-economic status, and that those who are socio-economically deprived are not only more likely to experience chronic pain compared to people with higher socioeconomic status, but they are also more likely to experience more severe pain and a greater level of pain-related disability^27, 28^. It is likely given the scale of use that lidocaine was prescribed off licence in Ireland for GMS patients with other types of neuropathic pain. GMS patients are less likely to have private health insurance and are largely dependent on the public healthcare system, and consequently the long waiting lists for these patients in accessing non-pharmacological therapies such as physical and psychological therapies as well as specialist pain clinics and surgery clinics may influence prescribing practices. The large difference in baseline rates may also have impacted on the implementation of the interventions. Although the appropriate rate of prescribing is unknown, it is likely that the scale of off-label or inappropriate use in Ireland was far greater than in England, with a much greater scope for reductions in prescribing. It is possible that had the scale of off-label lidocaine prescribing been similar in England, the introduction of the guidance may have had a more substantial effect.

A strength of this study is the use of robust pharmacy dispensing data. Data obtained from the OpenPrescribing platform relates to all prescriptions issued by general practices in England and dispensed in any community pharmacy in the UK. Similarly, PCRS data comprises all prescriptions dispensed under the GMS scheme. The use of real prescribing and dispensing data sourced from pharmacy claims minimises the potential for obtaining a biased sample, and additionally eliminates the possibility of recall bias. Data on dispensing reflects medications that a patient received, versus studies of prescribing data where a patient may not fill the prescription. The study does however have some limitations. While the English NHS data includes prescriptions issued to the whole population, the GMS scheme in Ireland is means-tested and therefore represents an older and lower socioeconomic subset of the Irish population. Additionally, the use of aggregate level data means that we were not able to examine whether increases in recommended alternatives (i.e., capsaicin) was actually among people who switched from lidocaine plasters. It also limits the ability to examine unintended consequences of the interventions, i.e., switching of patients to potentially riskier therapies.

More broadly, this study highlights the importance of open data. The NHS has published publicly-available GP prescribing data every month since 2011 for anyone to interrogate. This has supported a rich ecosystem of teams both inside and outside the NHS using differing tools and approaches to monitor data and give feedback to GPs to improve prescribing. Analysis conducted on this NHS open data has also supported original research on a substantial range of prescribing topics^19, 21, 29-32^,and data feedback to GPs has been shown to improve prescribing^33-36^. We strongly recommend that the HSE advances plans to publish similar data for GPs prescribing in Ireland in line with national open data polices^37^. This can support a similarly rich ecosystem of feedback to clinicians in Ireland and additionally our paper has demonstrated that by harnessing multiple countries data we can do innovative research into real world healthcare policy programmes.

## Conclusion

Our study has shown the effects of two different interventions aiming to decrease low-value prescribing and has demonstrated a more substantial reduction in prescribing in a high prescribing setting in Ireland where a patient approval system was introduced, compared to issuing guidance in England as a lower prescribing setting. More research on the effects of these interventions beyond prescribing rates and expenditure, including impact on patient outcomes, is warranted.

## Data Availability

Code and data for the analysis of NHS data is available from https://github.com/ebmdatalab/lidocaine-eng-ire. Code for the analysis of GMS data is available from https://zenodo.org/record/7287510#.Y-48eXbP02x. GMS data in aggregated form can be requested from the HSE PCRS at https://www.hse.ie/eng/staff/pcrs/pcrs-publications/.

## Contributors statement

BMK and FM conceived the study. MM, FB, CK, MF, EW, MEW, DC, TF, BMK and FM designed the study. MM, RC, SCJB, PI, DE, BG, BMK, and FM collected and curated the data. MM analysed the data and FM validated the data analysis. All authors were involved the interpretation of the analysis results. MM drafted the manuscript, and FB, CK, MF, EW, MEW, DC, TF, RC, SCJB, PI, DE, BG, BMK and FM critically revised the manuscript. FM acquired funding for the study.

## Declaration of interests

The authors declare no competing interests in relation to this study. All authors have completed the ICMJE (International Committee of Medical Journal Editors) uniform disclosure form at www.icmje.org/coi_disclosure.pdf and declare the following: MF and FM have received research funding from the Health Research Board in Ireland (HRB). BG has received research funding from the Laura and John Arnold Foundation, the NHS National Institute for Health Research (NIHR), the NIHR School of Primary Care Research, the NIHR Oxford Biomedical Research Centre, the Mohn-Westlake Foundation, NIHR Applied Research Collaboration Oxford and Thames Valley, Wellcome Trust, the Good Thinking Foundation, Health Data Research UK, the Health Foundation, the World Health Organization, UKRI, Asthma UK, the British Lung Foundation, and the Longitudinal Health and Wellbeing strand of the National Core Studies program; he also receives personal income from speaking and writing for lay audiences on the misuse of science. BMK is also employed by NHS England as a pharmacist adviser and supported development of NHS England guidance on lidocaine plasters. All other University of Oxford authors are employed on BG’s grants.

## Funding

This study is funded by the Health Research Board in Ireland (HRB) through the Secondary Data Analysis Projects scheme (CDRx project, PI FM, grant number SDAP-2019-023). The funder had no role in in study design; in the collection, analysis, and interpretation of data; in the writing of this paper; or in the decision to submit this paper for publication. EW is funded by a HRB Emerging Clinician Scientist Award (grant number: ECSA/2020/002). MEW is funded by a HRB Applying Research into Policy and Practice Award (ARPP/2020/004). OpenPrescribing.net is currently funded by NHS England’s Primary Care and Medicines Analytics Unit. All other Bennett funding information is available at https://www.bennett.ox.ac.uk/

## Notes

### Competing Interest Statement

The authors have declared no competing interest.

### Author Declarations

This study was approved by the RCSI University of Medicine and Health Sciences Human Research Ethics Committee (REC202201015).

