## Supplementary data for "The impact of lidocaine plaster prescribing reduction strategies: a comparison of two national health services in Europe"

**Supplementary materials**

**Supplementary table 1a. Quantity time-series results for Ireland**

| **Quantity of lidocaine patches per 1,000 GMS population** | **Coefficient** | **95% CI** | **P-value** |
| --- | --- | --- | --- |
| **Pre-Sep 2017 monthly trend** | 7.16 | 6.69 to 7.62 | <0.001 |
| **Sep 2017 immediate post intervention change** | -149.4 | -173.26 to -125.55 | <0.001 |
| **Sep 2017 – Nov 2017 trend change (versus pre-Sep 2017)** | -63.76 | -80.94 to -46.57 | <0.001 |
| **Dec 2017 immediate post intervention change** | -153.24 | -191.52 to -114.96 | <0.001 |
| **Dec 2017 – Dec 2019 trend change (versus pre-Sep 2017)** | 57.61 | 40.42 to 74.8 | <0.001 |
| **Jan 2015 dispensing rate (baseline)** | 267.09 | 236.74 to 297.43 | <0.001 |
| **Postintervention linear trends** |  |  |  |
| **Linear trend** | **Coefficient** | **95% CI** | **P-value** |
| Sep 2017 – Nov 2017 | -56.6 | -73.77 to -39.42 | <0.001 |
| Dec 2017 – Dec 2019 | 1.01 | 0.34 to 1.68 | 0.003 |

**Supplementary table 1b. Quantity time-series results for England**

| **Quantity of lidocaine patches per 1,000 NHS population** | **Coefficient** | **95% CI** | **P-value** |
| --- | --- | --- | --- |
| **Pre-Aug 2017 monthly trend** | 0.0961 | 0.0837 to 0.1085 | <0.001 |
| **Aug 2017 immediate post intervention change** | -0.6233 | -0.9429 to -0.3037 | <0.001 |
| **Aug 2017 – Dec 2019 trend change** | -0.1696 | -0.1881 to -0.1510 | <0.001 |
| **Jan 2015 dispensing rate (baseline)** | 10.8278 | 8.552 to 13.1035 | <0.001 |
| **Postintervention linear trends** |  |  |  |
| **Linear trend** | **Coefficient** | **95% CI** | **P-value** |
| Aug 2017 – Dec 2019 | -0.0735 | -0.0872 to -0.0597 | <0.001 |

**Supplementary table 2a. Cost time-series results for Ireland**

| **Total cost of lidocaine patches (€)** | **Coefficient** | **95% CI** | **P-value** |
| --- | --- | --- | --- |
| **Pre-Sep 2017 monthly trend** | 751.99 | 638.99 to 864.99 | <0.001 |
| **Sep 2017 immediate post intervention change** | -22,569.16 | -28,365.01 to -16,773.3 | <0.001 |
| **Sep 2017 – Nov 2017 trend change (versus pre-Sep 2017)** | -10,260.17 | -14,434.98 to -6,085.35 | <0.001 |
| **Dec 2017 immediate post intervention change** | -27,571.42 | -36,873.45 to-18,269.38 | <0.001 |
| **Dec 2017 – Dec 2019 trend change (versus pre-Sep 2017)** | 9,690.42 | 5,513.92 to 13,866.91 | <0.001 |
| **Jan 2015 dispensing rate (baseline)** | 58,457.23 | 50,876.34 to 66,038.11 | <0.001 |
| **Postintervention linear trends** |  |  |  |
| **Linear trend** | **Coefficient** | **95% CI** | **P-value** |
| Sep 2017 – Nov 2017 | -9,508.18 | -13,681.46 to -5,334.89 | <0.001 |
| Dec 2017 – Dec 2019 | 182.24 | 18.55 to 345.93 | 0.029 |

**Supplementary table 2b. Cost time-series results for England**

| **Total cost of lidocaine patches (£)** | **Coefficient** | **95% CI** | **P-value** |
| --- | --- | --- | --- |
| **Pre-Aug 2017 monthly trend** | 94.38 | 82.26 to 106.49 | <0.001 |
| **Aug 2017 immediate post intervention change** | -736.79 | -1,048.20 to -425.37 | <0.001 |
| **Aug 2017 – Dec 2019 trend change** | -196.48 | -214.54 to -178.43 | <0.001 |
| **Jan 2015 dispensing rate (baseline)** | 11,977.82 | 9,006.41 to 14,949.24 | <0.001 |
| **Postintervention linear trends** |  |  |  |
| **Linear trend** | **Coefficient** | **95% CI** | **P-value** |
| Aug 2017 – Dec 2019 | -102.11 | -115.5 to -88.72 | <0.001 |

 
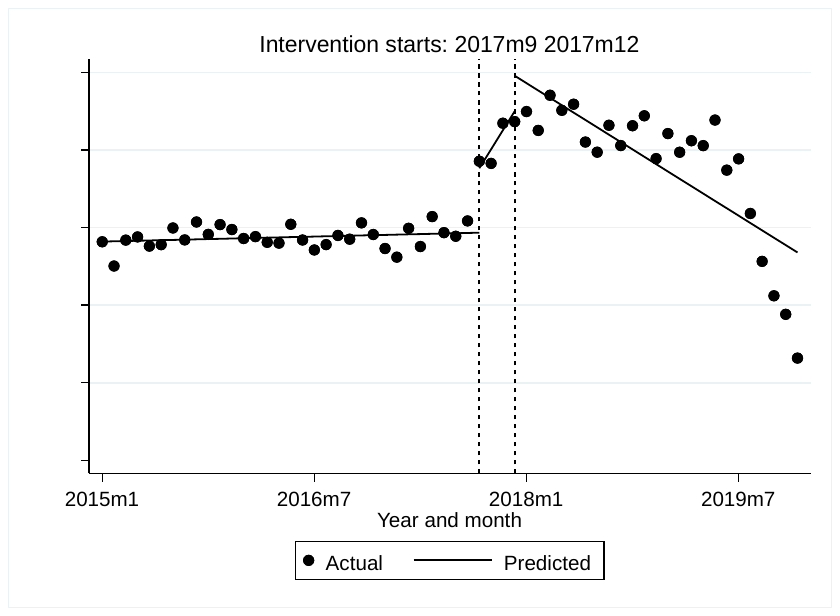

**Supplementary figure 1a. Time-series of dispensing rate of capsaicin per 1,000 for Ireland**

  
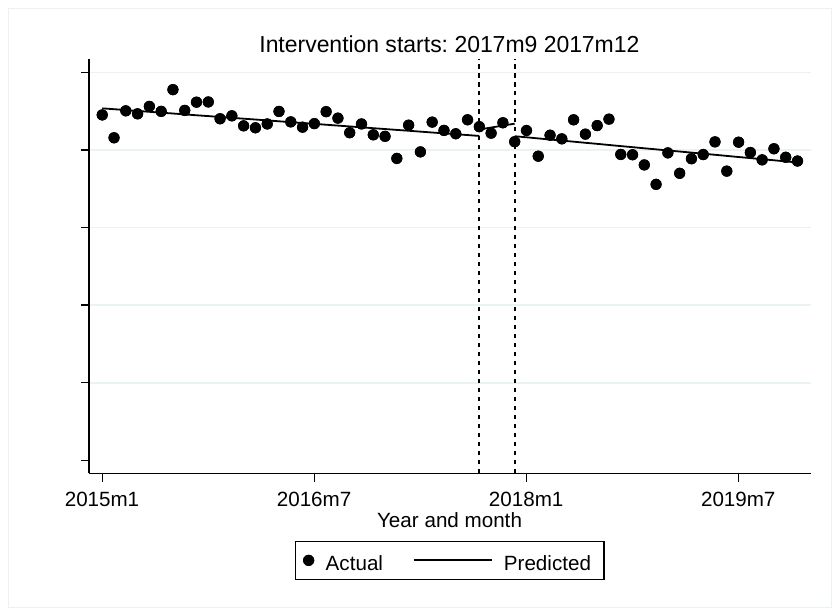

**Supplementary figure 2b. Time-series of dispensing rate of diclofenac per 1,000 for Ireland**

 
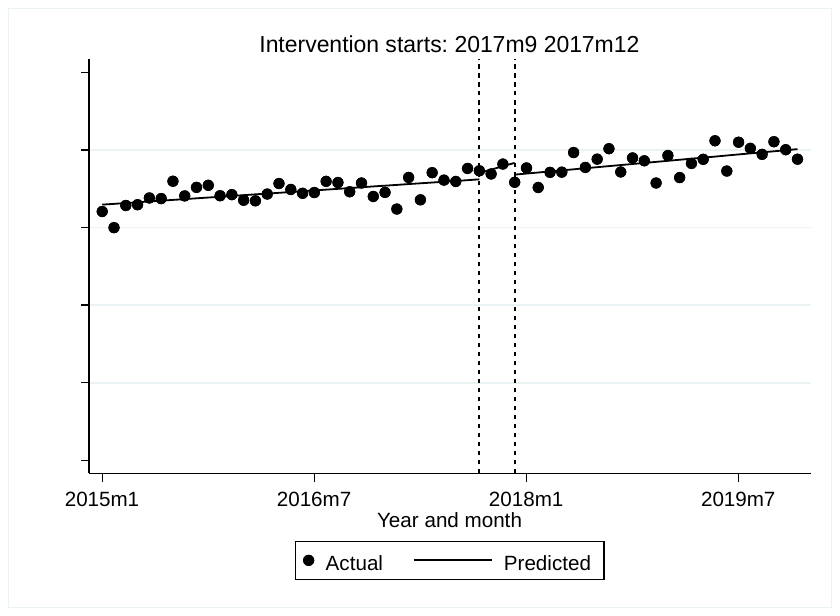

**Supplementary figure 1c. Time-series of dispensing rate of topical NSAIDs per 1,000 for Ireland**
